# Single-dose mRNA vaccine effectiveness against SARS-CoV-2 in healthcare workers extending 16 weeks post-vaccination: a test-negative design from Quebec, Canada

**DOI:** 10.1101/2021.07.19.21260445

**Authors:** Sara Carazo, Denis Talbot, Nicole Boulianne, Marc Brisson, Rodica Gilca, Geneviève Deceuninck, Nicholas Brousseau, Mélanie Drolet, Manale Ouakki, Chantal Sauvageau, Sapha Barkati, Elise Fortin, Alex Carignan, Philippe De Wals, Danuta M. Skowronski, Gaston De Serres

## Abstract

**Introduction:** In Canada, first and second doses of mRNA vaccines against SARS-CoV-2 were uniquely spaced 16 weeks apart, but the duration of single-dose protection remains uncertain. We estimated one- and two-dose mRNA vaccine effectiveness (VE) among healthcare workers (HCWs) in Quebec, Canada including protection against varying outcome severity, variants of concern (VOC), and the stability of single-dose protection out to 16 weeks post-vaccination.

**Methods:** A test-negative design compared vaccination among SARS-CoV-2 test-positive and weekly-matched (10:1), randomly-sampled, test-negative HCWs using linked surveillance and immunization databases. Vaccine status was defined by one dose ≥14 days or two doses ≥7 days before illness onset or specimen collection. Adjusted VE was estimated by conditional logistic regression.

**Results:** Primary analysis included 5,316 cases and 53,160 controls. Single-dose VE was 70% (95%CI: 68-73) against SARS-CoV-2 infection, 73% (95%CI: 71-75) against COVID-19 illness and 97% (95%CI: 92-99) against associated hospitalization. Two-dose VE was 86% (95%CI: 81-90) and 93% (95%CI: 89-95), respectively, with no associated hospitalizations. VE was higher for non-VOC than VOC (73% Alpha) among single-dose (77%, 95%CI: 73-81 versus 63%, 95%CI: 57-67) but not two-dose recipients (87%, 95%CI: 57-96 versus 94%, 95%CI: 89-96). Across 16 weeks, no decline in single-dose VE was observed with appropriate stratification based upon prioritized vaccination determined by higher versus lower likelihood of direct patient contact.

**Conclusion:** One mRNA vaccine dose provided substantial and sustained protection to HCWs extending at least four months post-vaccination. In circumstances of vaccine shortage, delaying the second dose may be a pertinent public health strategy to consider.

## INTRODUCTION

In December 2020, two mRNA vaccines against SARS-CoV-2 were authorized in Canada based upon a schedule of two doses spaced 3 weeks (BNT162b2 (Pfizer-BioNTech)) or 4 weeks (mRNA-1273 (Moderna)) apart [1]. Phase III randomized-controlled trials showed vaccine efficacy that exceeded 90% for both products beginning from 14 days after a single dose but did not inform protection beyond 3-4 weeks post-vaccination [2–5]. Healthcare workers (HCWs) were amongst the first prioritized for COVID-19 vaccination and several observational studies have since reported vaccine effectiveness (VE) after a single dose but most had short follow-up periods, the longest extending 8 weeks post-vaccination [6–10].

In the context of limited vaccine supply, the Quebec Immunization Committee (QIC) recommended that the province of Quebec, Canada defer the second dose of vaccine in order to optimize first-dose coverage and provide protection to as many high-risk individuals as possible against COVID-related hospitalizations and deaths [11]. The QIC did not pre-specify the interval for second-dose administration, relying upon real-time monitoring of single-dose VE and adaptation in the event of waning protection [12]. The vaccination campaign in Quebec began December 14, 2020, and initially targeted long-term care facility (LTCF) residents and healthcare workers (HCW) with direct patient contact. On March 3, 2021, the Canadian National Advisory Committee on Immunization and the Quebec Ministry of Health set the interval between doses at 16 weeks based upon expected vaccine supply, ethical considerations and short-term but reassuring VE findings [13,14]. Herein, we compare one- and two-dose mRNA VE against SARS-CoV-2, including varying outcome severity and variants of concern (VOC), among HCWs in Quebec and assess the stability of single-dose protection across 16 weeks post-vaccination.

## METHODS

### Study design

The study used a test-negative design: HCWs who tested RT-PCR positive for SARS-CoV-2 during the study period were cases; HCWs who tested RT-PCR negative were controls.

The case reference date was defined hierarchically as the date of symptom onset (83.5%) or if not available, then the date of specimen collection (16.5%). The control reference date was defined by specimen collection date. To account for time-varying likelihoods of SARS-CoV-2 exposure and vaccination [15], a density sampling approach was used with 10 randomly-sampled controls per case matched by week of reference date. A HCW could be sampled several times as a control over the study period (but only once per week) and could subsequently be included as a case. All cases were censored at their reference date.

### Population

The study population included publicly-paid HCWs but not privately-paid HCWs (in clinics, pharmacies, private seniors’ residences, etc.) or physicians who are paid by the provincial Medicare system.

Participants were excluded if: they were a confirmed SARS-CoV-2 case (RT-PCR confirmed or epidemiologically-linked) before January 17, 2021; were missing a unique personal identifying number (PIN) used for data linkage; had an invalid vaccination date (before December 14^th^); or were <18 or ≥75 years old. HCWs in child and youth protection centers or those hired temporarily for pandemic work by Ministerial order were also excluded. Finally, AstraZeneca vaccine recipients were excluded from the date of vaccination because their limited number precluded VE estimation.

### Data sources

Using the unique PIN, the cohort of all publicly-funded HCWs in the province was linked with: 1) the provincial database of all SARS-CoV-2 infections reported to public health since pandemic start, including associated clinical details collected during case investigation by public health authorities; 2) the administrative hospitalization database and the chronic disease surveillance system, which integrates information on pre-existing medical conditions; 3) the Quebec provincial immunization registry which is a census of all Quebec residents insured under the universal publicly-funded health care system, including whether or not vaccinated, vaccination date(s) and type of vaccine received; 4) the provincial centralized laboratory database, including the dates, results and reason for all RT-PCR tests for SARS-CoV-2 across the province; and 5) variant of concern (VOC) PCR screening assay results used to identify signature mutations (69-70 deletion, N501Y, E484K) to genetically characterize and categorize viruses. VOC screening was undertaken on a convenience sample of 10% of SARS-CoV-2 positive specimens in January 2021; on nearly 100% of specimens in February and March; and on about 85% of specimens from April 2021 onwards when the VOC prevalence exceeded 90% (Supplementary Figure 1) [16]. As of June 6, 2021, 89% of identified VOC cases were the Alpha variant (Pango lineage: B.1.1.7) [17].

The study period included HCWs with specimen reference date between January 17 (epidemiological week 3) and June 5 (week 22), 2021 (Figure 1), taking into account the immunization start-date and a several-week lag for vaccine effect. Data were extracted on June 17^th^ 2021, allowing additional 14-day lag to capture associated hospitalizations.

**Figure 1.**
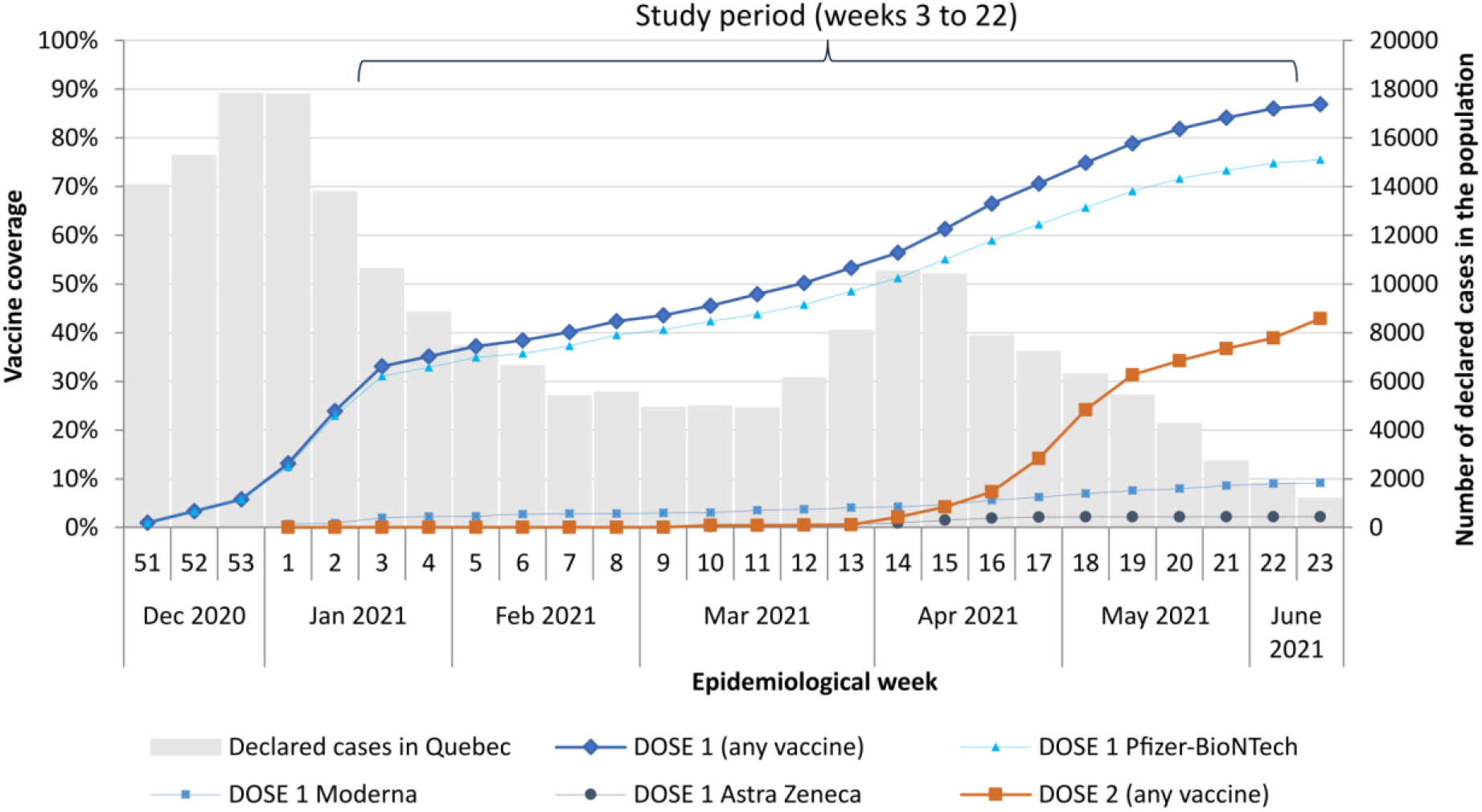
Vaccination coverage in the cohort of healthcare workers and total number of reported COVID-19 cases in the population per week, Quebec, Canada.

### Vaccination and outcome definitions

Vaccination status was defined in relation to the reference date. In primary analysis, a participant was deemed a single- or two-dose vaccinee if the doses were received ≥14 days or ≥7 days before the reference date (with day of vaccination being day 0), respectively, requiring ≥3 weeks between doses. HCWs who received no vaccine doses at any time on or before the reference date were considered unvaccinated whereas those who received the first dose <14 days or second dose <7 days prior were excluded. RT-PCR confirmed SARS-CoV-2 outcomes of varying severity were explored, including: any infection; symptomatic infection of any severity (specified hereafter as COVID-19); and COVID-19-related hospitalization (occurring within 30 days of illness onset).

### VE analysis

Odds ratios (OR) and their 95% confidence intervals (95%CI) among one- and two-dose vaccinees relative to unvaccinated HCWs were estimated by multivariate conditional logistic regression using the matching week as strata and adjusting for potential confounders. VE and 95% CIs were derived as: **(1 – *0 Radjusted*) x 100**.

Adjustment variables included: age group (18-29, 30-39, 40-49, 50-59, 60-74 years); sex; healthcare setting (hospitals and local community health centers, LTCFs, rehabilitation centers, home care, other); occupation (nurse, nursing assistant, personal healthcare support worker, other technical and health assisting occupations, administrative and management staff, healthcare technician, social workers, others); region of the healthcare setting (18 in Quebec); and presence of 17 possible comorbidities associated with increased COVID-19 hospitalization risk (known for ∼90% of HCWs) [18].

In addition to overall primary analyses by outcome severity, sensitivity analyses included: (a) stratifying by HCW priority group based upon those targeted before January 31 (week 5) or since February 21 (week 8) reflecting higher and lower frequency of direct patient contact and baseline infection risk, respectively; (b) stratifying by VOC status; (c) restricting to HCWs with data on 17 pre-existing conditions of comorbidity (at least one, at least two or by the number of 0, 1, 2, 3, 4, ≥5 conditions); and (d) stratifying by reason for testing (compatible symptoms, outbreak or systematic screening).

### Ethical aspects

This study was conducted as a surveillance and evaluation protocol with the legal mandate of the National Director of Public Health of Quebec and with the requirement for ethics approval thereby waived under the Public Health Act. It has been approved by the Research ethics board of the CHU de Québec-Université Laval.

## RESULTS

### Study population

Of the 342,138 HCWs in the initial cohort, 333,832 (97.6%) were successfully linked with the immunization registry. As of June 5 (end of week 22), 86.0% of individual HCWs in the cohort had received at least one vaccine dose (88.0% Pfizer-BioNTech, 9.4% Moderna, and 2.6% AstraZeneca vaccine) and 38.9% had received two doses (Figure 1).

We excluded: 8% of HCWs who were confirmed COVID-19 cases before January 17, 2021; 4% who worked in child and youth protection centers; 3% with missing PIN; 1% temporarily hired by Ministerial order; and <1% with an invalid vaccination date. Among the 284,637 remaining HCWs, 115,288 had no testing during the study period leaving 169,349 HCWs with 548,796 specimens for VE analysis. There were 5,316 cases with 53,160 controls randomly-sampled among negative tests (Supplementary Figure S1), of which 10.8% cases and 11.8% controls vaccinated 0-13 days or 0-6 days before the first or second dose, respectively, were excluded from primary analyses.

### Characteristics of cases and controls

At their reference date, 23.8% of cases and 49.1% of controls had received one dose ≥14 days earlier and 0.9% of cases and 3.9% of controls had received two doses ≥7 days earlier. The percentage of controls vaccinated with at least one dose ≥14 days before the reference date increased with age, from 49.1% in 18-29-year-olds to 61.5% among 60-74-year-olds (Supplementary Figure S2). For those who received 2 doses, the median interval between doses was nearly 16 weeks (111 days for cases and 112 days for controls). The median follow-up time for one-dose vaccinees was 56 days and for two-dose vaccinees was 18 days. Among all cases, 80.5% had COVID-19-related symptoms, 1.7% were hospitalized and one unvaccinated HCW died (Table 1). For symptomatic COVID-19 cases, the median and mean interval between symptom onset and testing was 1.0 and 1.3 days, respectively.

**Table 1.** Vaccination status, demographic and employment characteristics of cases and controls

|  | CASES |  |  |  |  |  | CONTROLS |  |
| --- | --- | --- | --- | --- | --- | --- | --- | --- |
|  | Any SARS-COV-2 |  | COVID-19 |  | Hospitalizations |  |  |  |
|  | infections |  |  |  |  |  |  |  |
|  | N=5316 |  | N=4277 |  | N=92 |  | N=53160 |  |
|  | N | % | N | % | N | % | N | % |
| **Vaccination status** (days before illness onset/ testing) |  |  |  |  |  |  |  |  |
| One-dose vaccinees (≥14 days) | 1266 | 23.8 | 970 | 22.7 | 4 | 4.4 | 26118 | 49.1 |
| *PFIZER-BioNTech vaccine* | 1202 | 22.6 | 924 | 21.6 | 4 | 4.4 | 24589 | 46.3 |
| *MODERNA vaccine* | 64 | 1.2 | 46 | 1.1 | 0 | 0.0 | 1529 | 2.9 |
| Two-dose vaccinees (≥7 days) | 50 | 0.9 | 20 | 0.5 | 0 | 0.0 | 2081 | 3.9 |
| *PFIZER-BioNTech vaccine* | 48 | 0.9 | 20 | 0.5 | 0 | 0.0 | 1954 | 3.7 |
| *MODERNA vaccine* | 2 | 0.0 | 0 | 0.0 | 0 | 0.0 | 126 | 0.2 |
| Vaccinated with at least 1 dose (≥14 days) <sup>a</sup> | 1373 | 25.8 | 1029 | 24.1 | 4 | 4.4 | 29201 | 54.9 |
| Unvaccinated | 3424 | 64.4 | 2813 | 65.8 | 86<sup>b</sup> | 93.5 | 18663 | 35.1 |
| Delay between doses in days (median and IQR) | 111 | 106–112 | 112 | 109–114 | NA |  | 112 | 107–112 |
| **Reasons for testing** |  |  |  |  |  |  |  |  |
| Symptoms compatible with COVID-19 | 2751 | 51.8 | 2619 | 61.2 | 44 | 47.8 | 8094 | 15.2 |
| Testing in the presence of an outbreak | 629 | 11.8 | 407 | 9.5 | 6 | 6.5 | 12517 | 23.6 |
| Systematic screening (asymptomatic) | 653 | 12.3 | 373 | 8.7 | 7 | 7.6 | 23940 | 45.0 |
| Other reason or unknown | 1283 | 24.1 | 878 | 20.5 | 35 | 38.0 | 8609 | 16.2 |
| **Variants of concern** (among 2889 positive specimens) |  |  |  |  |  |  |  |  |
| Any VOC | 1620 | 56.1 | 1368 | 58.1 | 43 | 67.2 | NA |  |
| Alpha variant (lineage B.1.1.7) | 1182 | 40.9 | 1003 | 42.6 | 31 | 48.4 | NA |  |
| VOC not detected | 944 | 32.7 | 784 | 33.3 | 15 | 23.4 | NA |  |
| Uninterpretable result | 325 | 11.2 | 203 | 8.6 | 6 | 9.4 | NA |  |
| **Demographic, clinical and employment characteristics** |  |  |  |  |  |  |  |  |
| Age (median and IQR) | 39 | 30–49 | 39 | 30–49 | 48.5 | 38–56 | 41 | 31–51 |
| 18-29 years | 1295 | 24.4 | 1049 | 24.5 | 9 | 9.8 | 10826 | 20.4 |
| 30-39 years | 1403 | 26.4 | 1157 | 27.1 | 17 | 18.5 | 13642 | 25.7 |
| 40-49 years | 1343 | 25.3 | 1067 | 25.0 | 24 | 26.1 | 13569 | 25.5 |
| 50-59 years | 1049 | 19.7 | 828 | 19.4 | 31 | 33.7 | 11957 | 22.5 |
| 60-74 years | 226 | 4.3 | 176 | 4.1 | 11 | 12.0 | 3166 | 6.0 |
| Sex, female | 4233 | 79.6 | 3459 | 80.9 | 69 | 75.0 | 44280 | 83.3 |
| Comorbidity (n=53075; 90.8%) <sup>c</sup> |  |  |  |  |  |  |  |  |
| At least one medical condition | 1617 | 30.8 | 1302 | 30.6 | 49 | 53.3 | 15963 | 33.1 |
| At least two medical conditions | 502 | 9.5 | 415 | 9.8 | 21 | 22.8 | 4845 | 10.1 |
| **Job category** |  |  |  |  |  |  |  |  |
| Nurse | 975 | 18.3 | 801 | 18.7 | 16 | 17.4 | 12519 | 23.6 |
| Nursing assistant | 474 | 8.9 | 372 | 8.7 | 12 | 13.0 | 4516 | 8.5 |
| Healthcare support worker | 1108 | 20.8 | 817 | 19.1 | 29 | 1.1 | 12357 | 23.2 |
| Other health assisting occupations | 662 | 12.5 | 503 | 11.8 | 10 | 31.5 | 5430 | 10.2 |
| Technical assisting occupations | 424 | 8.0 | 337 | 7.9 | 3 | 10.9 | 3154 | 5.9 |
| Administrative and management staff | 849 | 16.0 | 739 | 17.3 | 15 | 3.3 | 5305 | 10.0 |
| Healthcare technician | 349 | 6.6 | 289 | 6.8 | 4 | 16.3 | 5084 | 9.6 |
| Social worker | 402 | 7.6 | 360 | 8.4 | 2 | 4.4 | 3200 | 6.0 |
| Other | 73 | 1.4 | 59 | 1.4 | 1 | 2.2 | 1595 | 3.0 |
| **Facility** |  |  |  |  |  |  |  |  |
| Hospitals / local community health centers | 2857 | 53.7 | 2348 | 54.9 | 54 | 58.7 | 28563 | 53.7 |
| Long term health facilities | 1063 | 20.0 | 772 | 18.1 | 21 | 22.8 | 14890 | 28.0 |
| Rehabilitation centers | 283 | 5.3 | 229 | 5.4 | 1 | 1.1 | 1353 | 2.6 |
| Home care | 283 | 5.3 | 235 | 5.5 | 1 | 1.1 | 3029 | 5.7 |
| Other | 830 | 15.6 | 693 | 16.2 | 15 | 16.3 | 5325 | 10.0 |

<sup>a</sup> Including those vaccinated with 2 doses less than 7 days before the reference date
<sup>b</sup> 2 hospitalized cases were vaccinated 0-13 days before illness onset
<sup>c</sup> 17 medical conditions considered: hypertension, cardiovascular disease, neurological disorder, anemia, respiratory disease, diabetes, hypothyroidism, fluid and electrolyte disorder, cancer, kidney disease, obesity, psychosis, liver disease, immune disease, coagulopathy and paralysis
Abbreviations: IQR=Interquartile range; NA=not applicable; VOC=Variant of concern

Screening PCR tests to identify VOC were performed on 2889 (54.4%) positive specimens (91.5% during weeks 8 to 20): 1620 (56.1%) were a VOC and among them 73.0% were identified as the Alpha variant (Table 1).

Demographic and employment characteristics are provided in detail in Table 1.

### Vaccine effectiveness

#### Overall, by outcome severity

The overall adjusted single-dose mRNA VE was 70.4% (95%CI: 68.2-72.5) against any SARS-CoV-2 infection, 72.9% (95%CI: 70.6-75.0) against COVID-19 and 97.2% (95%CI: 92.3-99.0) against COVID-19-related hospitalization (Table 2). The overall adjusted two-dose mRNA VE was 85.8% (95%CI: 81.0-89.50) and 92.7% (95%CI: 88.5-95.4), respectively, with no associated hospitalizations. No differences were found by vaccine type (Pfizer-BioNTech or Moderna) (Table 2) or by age group (Supplementary Figure S2).

**Table 2.**
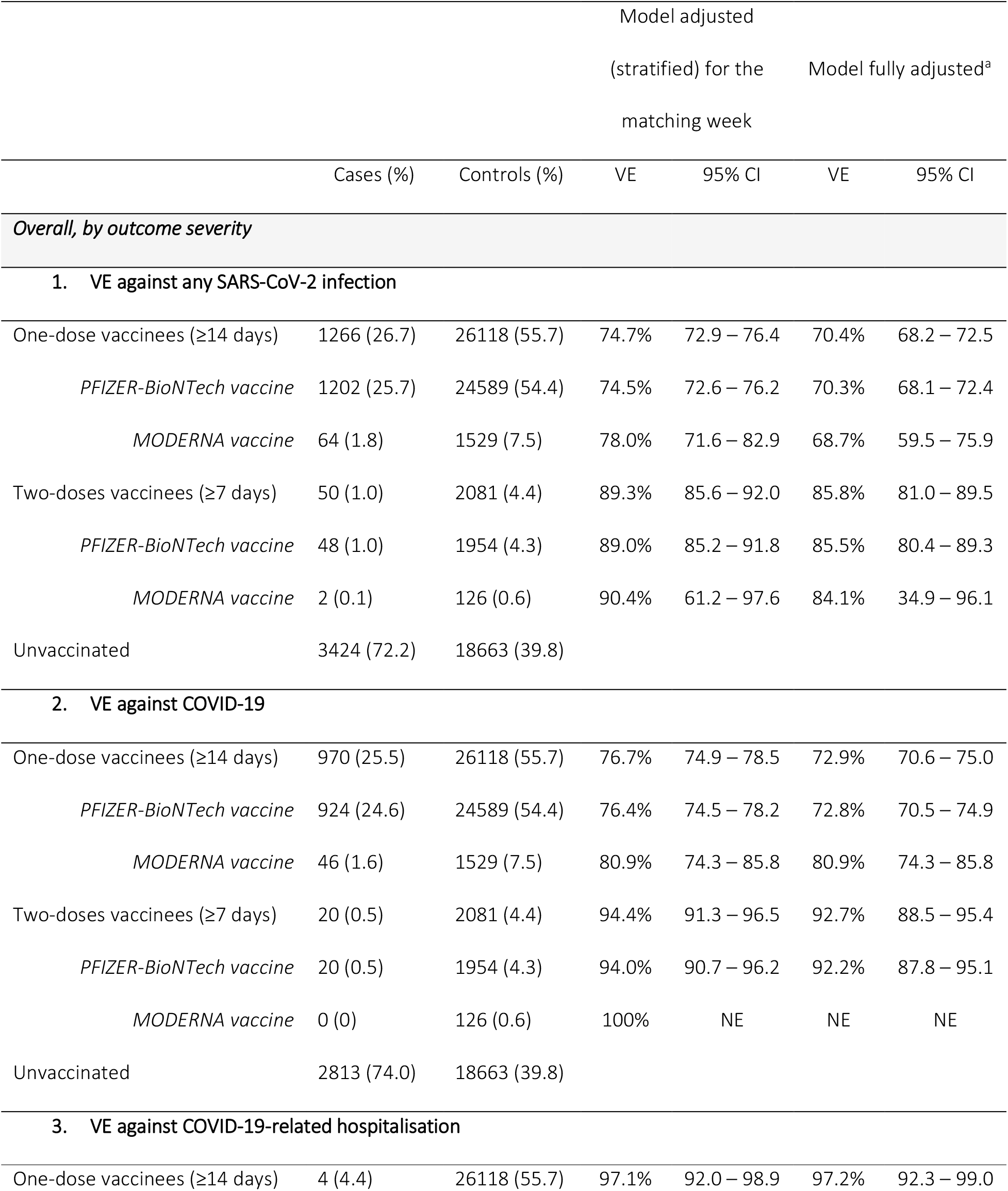
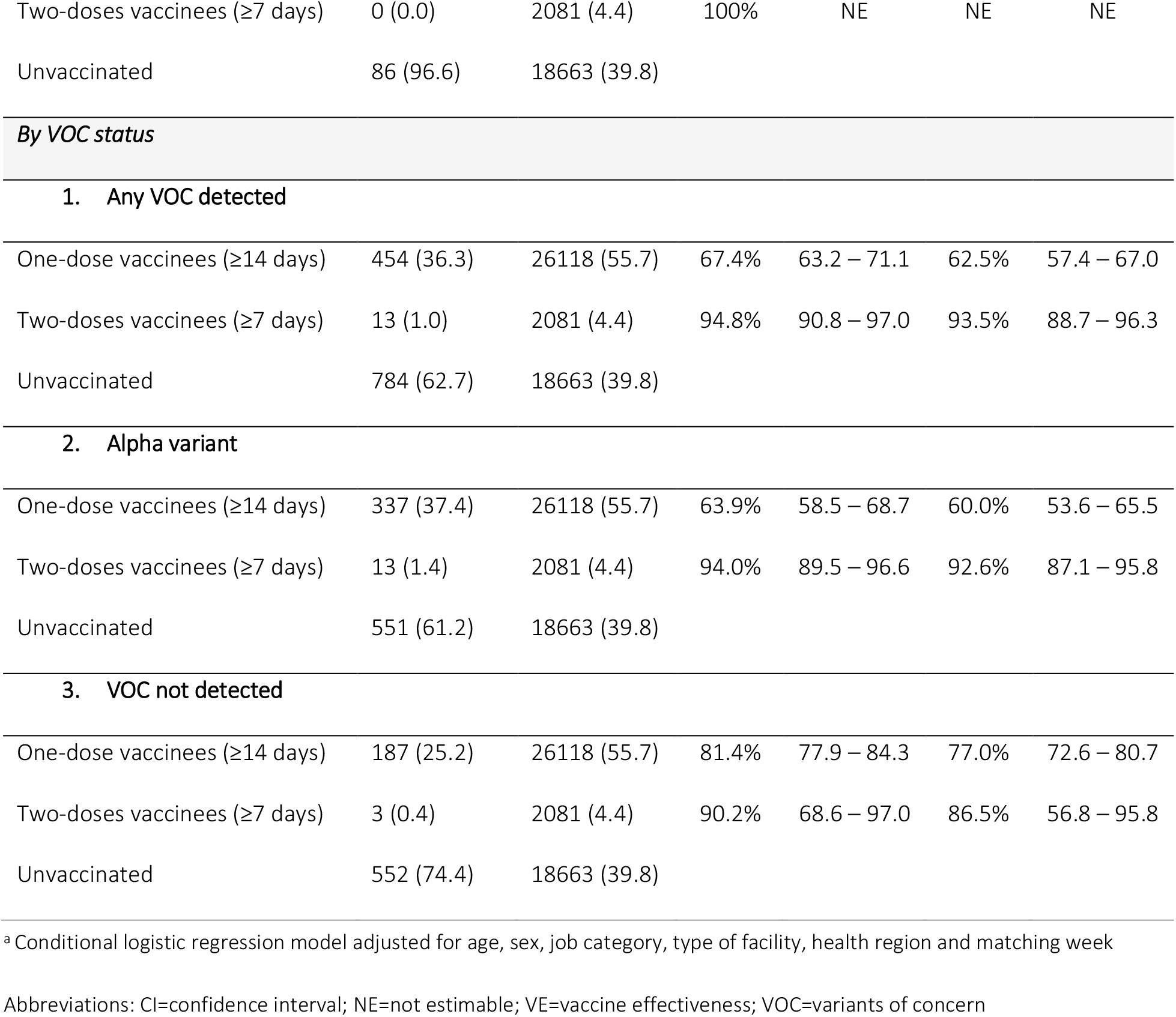
Overall vaccine effectiveness by outcome severity (SARS-CoV-2 infection, COVID-19 and COVID-19-related hospitalization) and by variant of concern status

One-dose VE against COVID-19 was 76-78% between 2- and 7-weeks post-vaccination, declining slightly to about 70% between 9- and 16-weeks post-vaccination (<1% were vaccinated >16 weeks prior) (Figure 2). Follow-up after two doses was too short for corresponding interval analyses.

**Figure 2.**
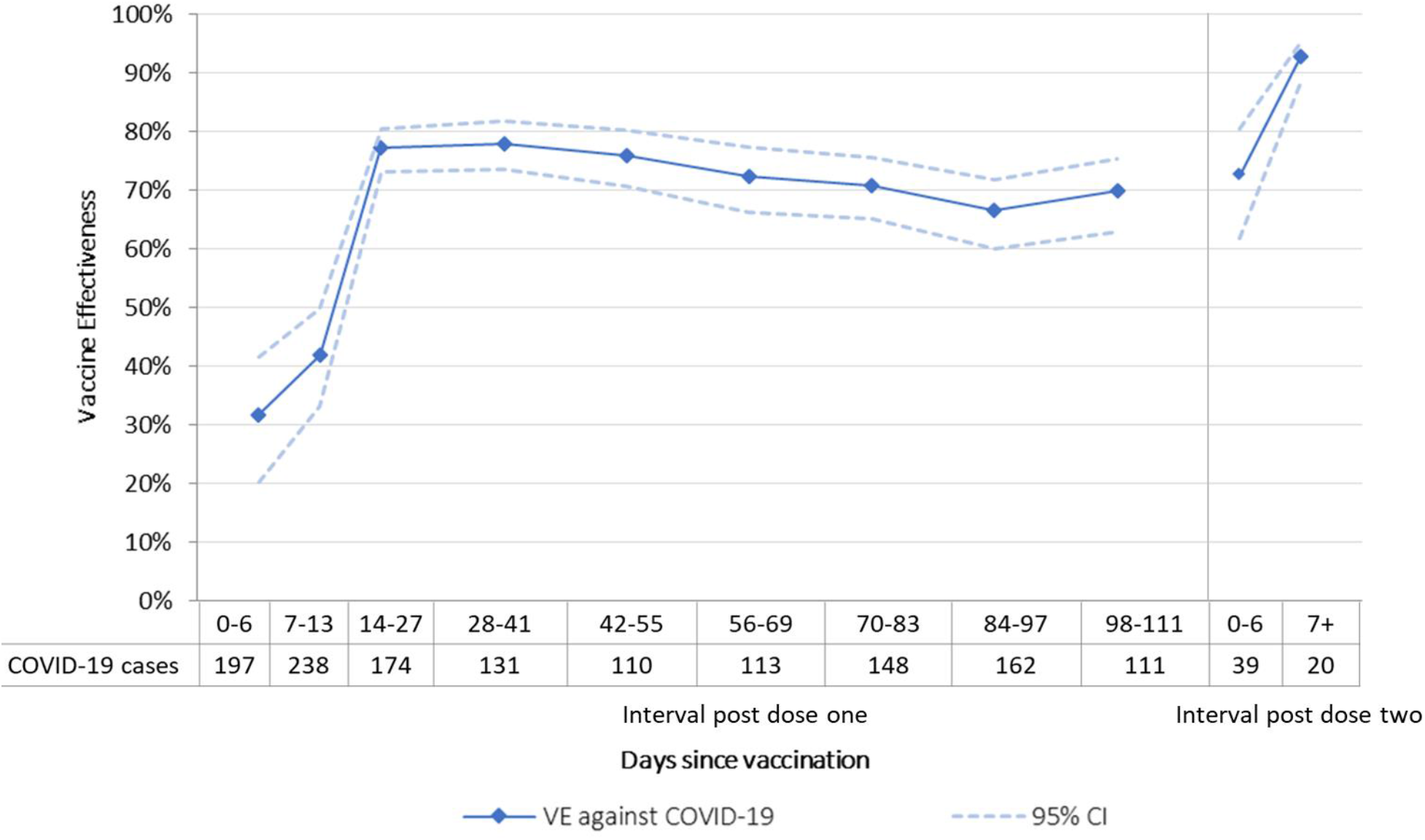
Vaccine effectiveness against COVID-19 by interval since vaccination Abbreviations: VE=Vaccine effectiveness; CI=Confidence Interval

#### By HCW priority group

Among HCWs first targeted for vaccination before week 5 (because of highest likelihood of direct patient contact and baseline infection risk), VE during the period 0-13 days after the first dose (when no vaccine effect is expected) was highly negative (−101.6%, 95%CI: −139.9 to −69.5).

Conversely, among HCWs first vaccinated after week 8 (at lower baseline infection risk), VE during the period 0-13 days was significantly higher at 43.7% (95%CI: 31.9-53.5). Thereafter, the earlier vs. later targeted HCWs had lower VE overall across the 2-16-week analysis period (52.2%, 95%CI: 47.1-56.9 vs 77.4%, 95%CI: 73.0-81.1), with neither target group showing decline in protection over that extended follow-up period (Supplementary Table S1 and Figure 3).

**Figure 3.**
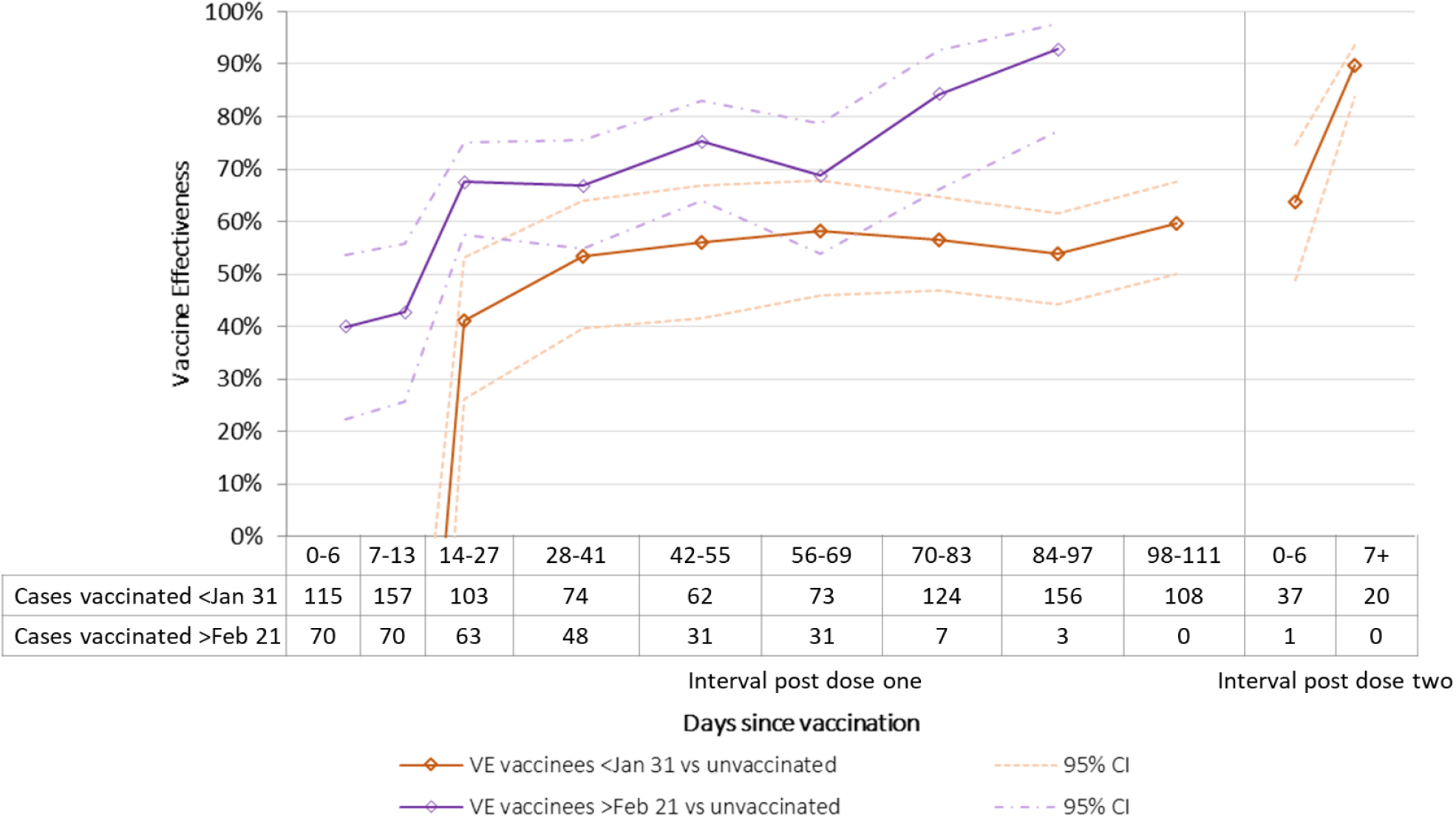
Vaccine effectiveness against COVID-19 in healthcare workers vaccinated before January 31^st^ 2021 (highest contacts with patients) and those vaccinated after February 20^th^ 2021 (fewer contacts with patients) by interval since vaccination Abbreviations: VE=Vaccine effectiveness; CI=Confidence Interval

#### By VOC status

With restriction to cases with screening VOC results, VE against COVID-19 was higher for non-VOC than VOC among single-dose (77.0%, 95%CI: 72.6-80.7 versus 62.5%, 95%CI: 57.4-67.0) but not two-dose recipients (86.5%, 95%CI: 56.8-95.8 versus 93.5%, 95%CI: 88.7-96.3). The Alpha-specific VE did not differ from VE against any VOC (including those with undetermined lineage) (Table 2) but was consistently lower than for non-VOC across the entire follow-up period (Figure 4). Among 90 hospitalized cases, 63 were analysed for VOC with the only vaccinated case bearing the Alpha variant.

**Figure 4.**
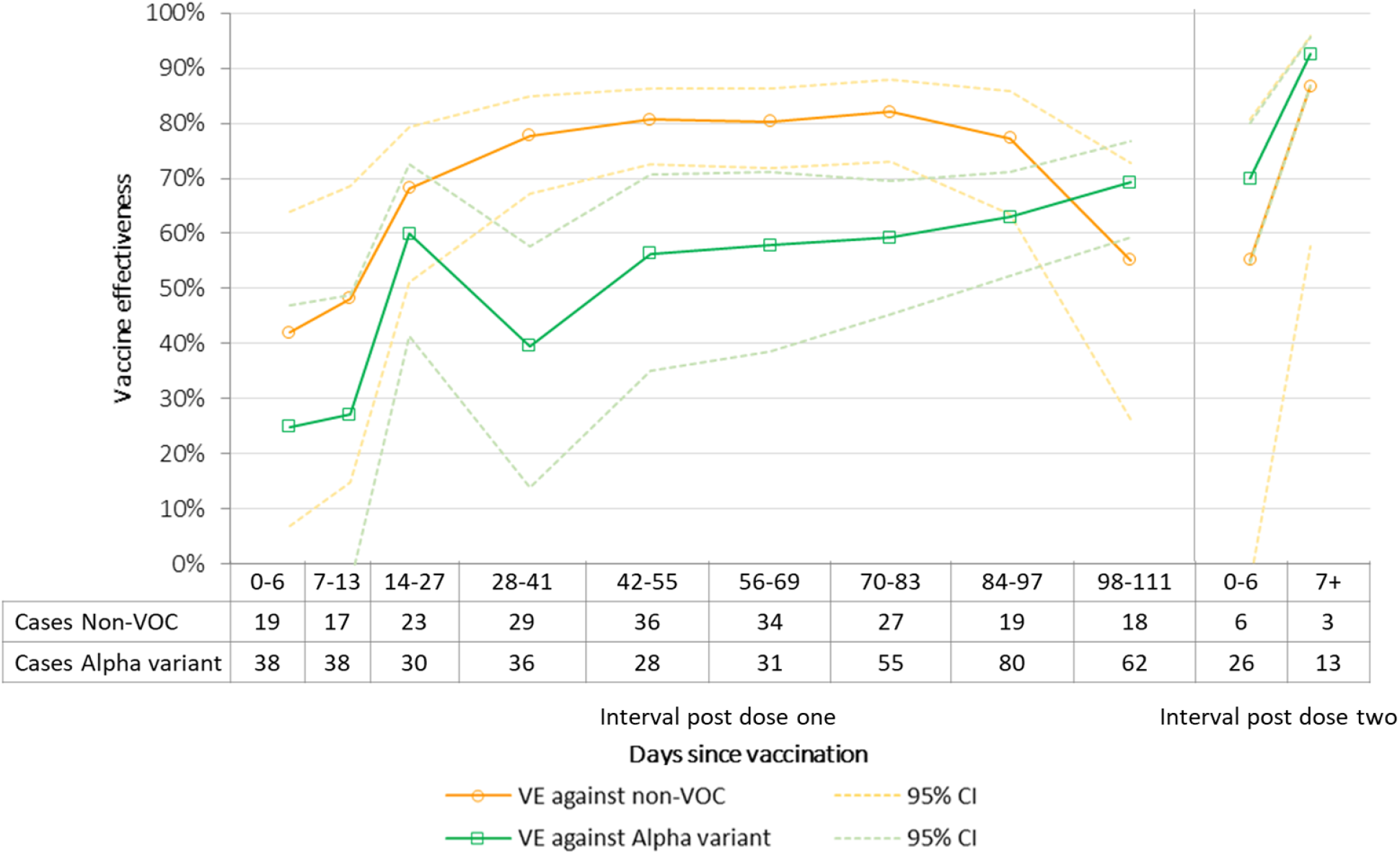
Vaccine effectiveness against COVID-19 by variant of concern (VOC) status (non-VOC or Alpha variant) by interval since vaccination Abbreviations: VE=Vaccine effectiveness; VOC=Variants of concern; CI=Confidence Interval

#### By comorbidity and testing indication

Adjustment for the presence of comorbidity did not meaningfully affect VE estimates (data not shown). In analyses stratified by testing indication, one-dose VE was lower for COVID-19 compatible symptoms (62.7%, 95%CI: 58.6-66.4) versus outbreak-(73.0%, 95%CI: 64.7-79.4) or screening-related testing (73.1%, 95%CI: 64.9-79.4). Two-dose estimates were higher for HCWs tested for symptomatic illness but with overlapping 95%CIs (Supplementary Table S1).

## DISCUSSION

In this observational study, we report single-dose mRNA VE of 70% against any SARS-CoV-2 infection and 73% against COVID-19 illness among HCWs in Quebec, Canada. Although VE was higher at 86% and 93%, respectively, after a second dose, VE against COVID-19 hospitalization was comparably high at >95% for both one- and two-dose recipients. Importantly, we provide the longest single-dose vaccination follow-up to date, showing substantial protection maintained for at least 16 weeks after receipt of just one dose of mRNA vaccine. Overall, our findings reinforce the recommendation for second-dose deferral and show that the interval between doses can be extended to at least four months where indicated due to scarce vaccine supply.

Other observational studies from the US, Israel and Europe have reported comparable Pfizer-BioNTech VE among HCWs beginning 14 and 7 days after dose one or two, respectively, but these involved only short follow-up periods. In two US studies, single-dose VE among HCWs, vaccinated according to the manufacturer’s schedule, was 78% against any infection [6], and 74% against symptomatic infection [7], with two dose VE of 97% and 94%, respectively [6,7]. Similarly, in an Israeli HCW cohort, single-dose VE was 75% against any SARS-CoV-2 infection from 15-28 days post-vaccination [9]. In Italy, single-dose VE between 14-21 days post-vaccination in HCWs was 83% against symptomatic infection and lower at 66% for ≥21 days without specification of the longest duration of follow-up [8]. In the UK, where the second dose was also deferred [19], the SIREN study reported VE against any SARS-CoV-2 infection of 72% from 21 to 69 days post-vaccination in HCWs systematically tested over a maximum of 8 weeks [10].

In our study, single-dose mRNA VE against hospitalization among HCWs was 97% across a 16-week period. Although evidence elsewhere supports high two-dose protection against hospitalization [20–22], few studies to date have reported single-dose protection and none over such an extended follow up period. In population-based studies, one-dose VE against hospitalizations among mostly older adults was 74% (14-20 days post-vaccination) in Israel, 77% (>14 days post-vaccination) in the US and 91% (14-34 days post-vaccination) in Scotland [21–23].

The Alpha (B.1.1.7) variant was the most prevalent SARS-CoV-2 virus in circulation in Quebec between late March and the end of our study period. The lower single-dose VE of 60% (95%CI: 54% to 66%) we report against symptomatic Alpha infection among HCWs is consistent with the lower single-dose VE of 67% against Alpha infection recently reported for older adults ≥70 years old from the province of British Columbia, Canada also based upon test-negative design [24]. Our higher two-dose estimate of 93% (95%CI: 87% to 96%) is also consistent with estimates exceeding 90% from Israel [20] and Qatar [25]. Of note, the later predominant contribution by and lower VE against the Alpha variant, whose prevalence among SARS-CoV-2 infections was <30% in mid-March (week 12) but increased to >90% at the end of the study, may have contributed to an apparent but perhaps artefactual decline in overall single-dose protection across the analysis period.

Furthermore, with respect to the question of potential waning of single-dose vaccine protection, we highlight an important methodological issue, critical for other investigators to consider. In particular, we illustrate the impact that confounding by indication can have when averaging VE across sub-groups with different exposure risks and who are sequentially prioritized or targeted for vaccination on that basis. HCWs earliest prioritized for vaccination because of highest baseline infection risk will also contribute most to the longest post-vaccination analysis intervals. In pooled analysis, their systematically higher infection risk and lower VE will lead to an overall, but erroneous, impression of declining single-dose VE generally with time since vaccination.

Conversely, with appropriate stratification based upon underlying differential in exposure and infection risk, we show single-dose VE to have been stable across the analysis period including both earlier and later prioritized HCW sub-sets. Properly addressing that methodological bias, we demonstrate no evidence for decrease in single-dose VE across a four-month follow-up period.

This study has limitations, foremost related to its observational design, subject to bias and confounding, and reliance on surveillance data subject to misclassification and missing information. Like others we could not fully adjust for differential exposure risk or fully ascertain the symptom profile notably after specimen testing. Despite easy access to testing, some asymptomatic infections were likely missed. HCWs with undetected infections before the study period could not be excluded leading to bias due to undiagnosed cases among vaccinated (overestimation) or unvaccinated (underestimation) participants [26]. A Ministerial order issued on April 9 requiring unvaccinated HCWs to be systematically tested every 3 days [27] may have increased detection of asymptomatic infections in unvaccinated individuals potentially leading to overestimation of VE against any infection at the end of the follow-up period but without affecting VE against COVID-19 or hospitalization. HCWs are active and relatively young adults and these results may not apply to older adults [28,29]. Even if adjustment for comorbidities did not change VE estimates, HCWs with medical conditions at high risk of severe disease were frequently removed from direct patient care duties during the pandemic and their VE might be lower than the estimates for all HCWs [22]. Despite these limitations, our study has strengths including its extended post-single-dose follow-up of a large and well-defined cohort and its several sensitivity and stratified analyses to address confounding due to time-dependent variables (vaccination prioritization and exposure risk) and variation in VOC circulation. Whereas VE estimates from our observational design may not precisely mimic short-term RCT estimates, the stable pattern of persistent single-dose protection we report across several months of follow-up is a unique and informative advantage over prior studies and may be the most meaningful with respect to public health implications for other areas still grappling with vaccine shortage.

In conclusion, one dose of mRNA vaccine reduced the risk of COVID-19 among HCWs by at least three-quarters (preventing three out of four symptomatic infections) and the associated risk of hospitalization by more than 95%, with such single-dose protection extending at least 16 weeks post-vaccination. Our findings of substantial and sustained single-dose VE, including against the Alpha variant, reinforce the option to defer the second dose of mRNA vaccine in circumstances of scarce vaccine supply and where broad single-dose coverage is critically needed.

## Data Availability

Data is not publicly available

## Funding

This work was supported by the Ministère de la santé et des services sociaux du Québec.

## Potential conflicts of interest

DT is supported by a career award from the Fonds de recherche du Québec – Santé. GDS received a grant from Pfizer for anti-meningococcal immunogenicity study.

## Acknowledgments

We would like to thank Louis Rochette for his work on preparing the healthcare workers’ dataset.

## Notes

### Competing Interest Statement

DT is supported by a career award from the Fonds de recherche du Quebec Sante. GDS received a grant from Pfizer for anti-meningococcal immunogenicity study. Other authors declared no competing interest.

### Funding Statement

This work was supported by the Ministere de la sante et des services sociaux du Quebec.

### Author Declarations

This study was conducted as a surveillance and evaluation protocol with the legal mandate of the National Director of Public Health of Quebec and with the requirement for ethics approval thereby waived under the Public Health Act. It has been approved by the Research ethics board of the CHU de Quebec-Universite Laval.

