## Supplemental material for "Single-dose mRNA vaccine effectiveness against SARS-CoV-2 in healthcare workers extending 16 weeks post-vaccination: a test-negative design from Quebec, Canada"

**Supplementary Figure S1**. Population flow chart

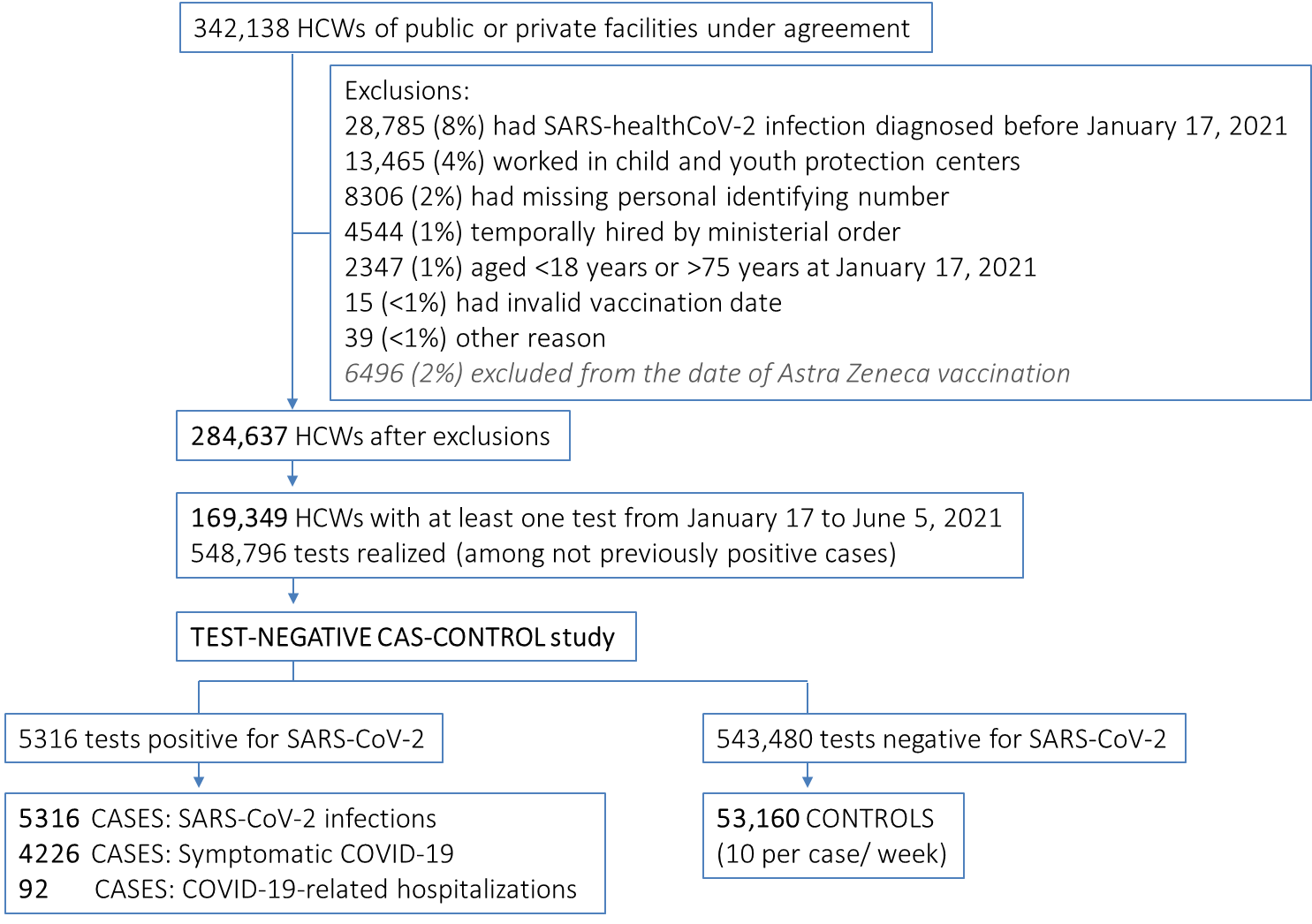

**Supplementary Figure S2**. Proportion of controls vaccinated with one and two doses ≥14 and ≥7 days before the reference date and adjusted vaccine effectiveness against COVID-19 by age group

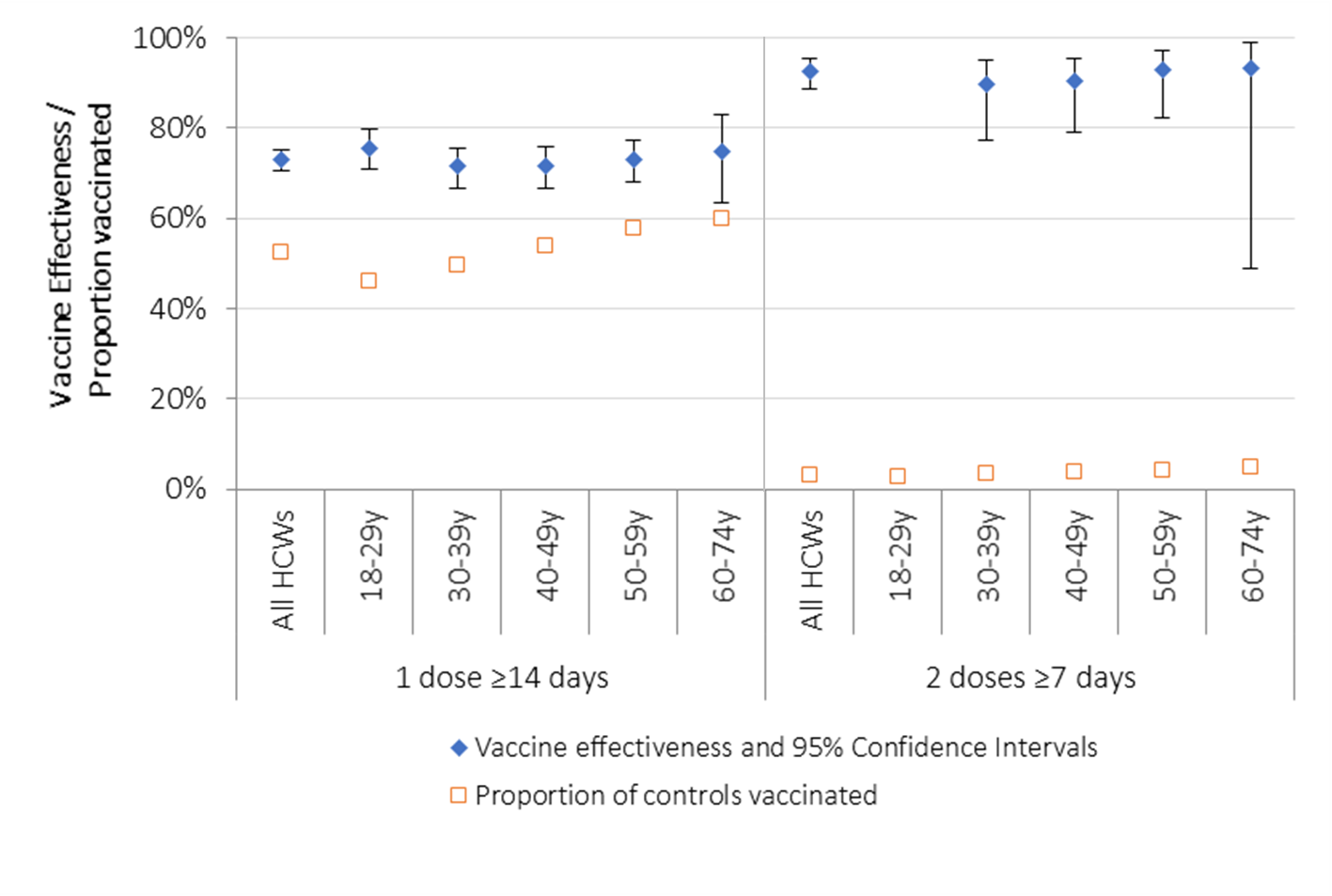

**Supplementary Figure S3**. Weekly cases of SARS-CoV-2 infection among healthcare workers included in the study and number of cases analyzed to identify variants of concern (VOC): Alpha variant, other or undetermined VOC and non-VOC

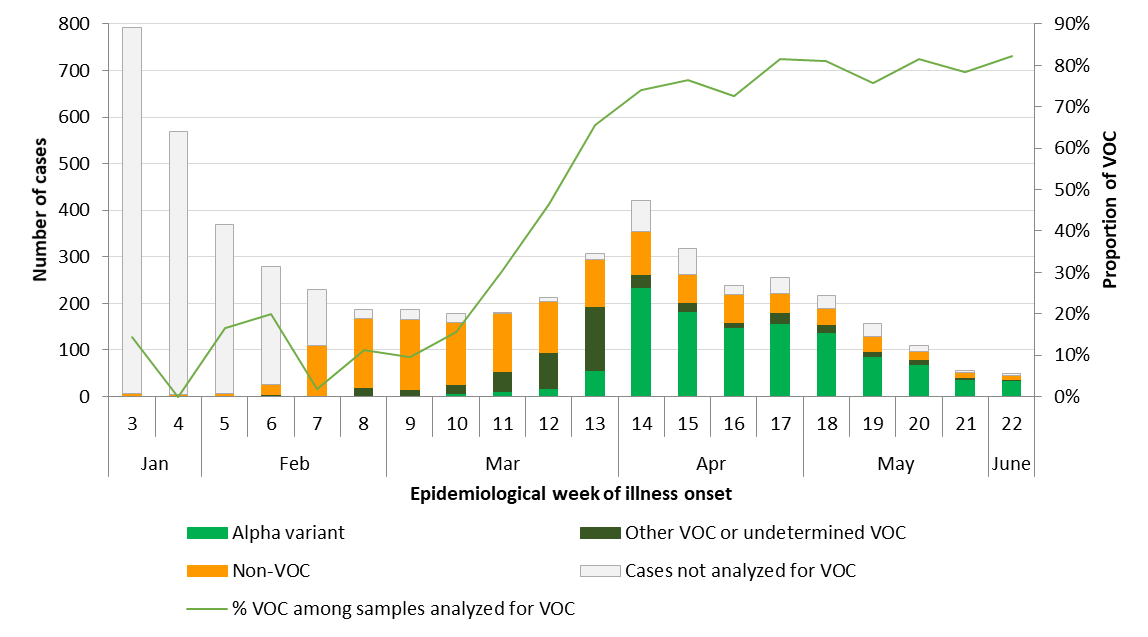

**Supplementary Table 1**: Sensitivity analyses of VE against COVID-19 (symptomatic infection) by priority group (vaccinated before January 31^st^ 2021 with highest contacts with patients and vaccinated after February 20^th^ 2021 with fewer contacts with patients) and reason for testing

|  | Cases (%) | Controls (%) | Model adjusted for matching week | | Model fully adjusted ^a^ | |
| --- | --- | --- | --- | --- | --- | --- |
|  |  |  | VE | 95% CI | VE | 95% CI |
| **By priority group:** |  |  |  |  |  |  |
| **Vaccinated Dec 14 to Jan 30 /Unvacc.**  Cases from Jan-17 to June-05 | 3134 (2484) | 31340 |  |  |  |  |
| One-dose vaccinees (≥14 days) | 721 (32.2) | 14551 (55.7) | 59.9% | 55.8 – 63.5 | 52.2% | 47.1 – 56.9 |
| Two-doses vaccinees (≥7 days) | 20 (0.9) | 1825 (4.4) | 92.2% | 87.7 – 95.1 | 89.4% | 83.2 – 93.3 |
| Unvaccinated | 1434 (65.9) | 12108 (39.8) |  |  |  |  |
| **Vaccinated Feb 21 to June 5 /Unvacc.**  Cases from Mars-07 to June-05 | 1902 (1559) | 19020 |  |  |  |  |
| One-dose vaccinees (≥14 days) | 183 (12.9) | 5677 (34.0) | 78.7% | 74.7 – 82.1 | 77.4% | 73.0 – 81.1 |
| Two-doses vaccinees (≥7 days) | 0 (0.0) | 736 (4.4) | 100% | NE | NE | NE |
| Unvaccinated | 1235 (87.1) | 10310 (61.7) |  |  |  |  |
| **By reason for testing:** |  |  |  |  |  |  |
| **Presence of COVID-19 symptoms** | 2744 (2744) | 27440 |  |  |  |  |
| One-dose vaccinees (≥14 days) | 615 (25.2) | 10586 (43.8) | 62.8% | 58.9 – 66.3 | 62.7% | 58.6 – 66.4 |
| Two-doses vaccinees (≥7 days) | 8 (0.3) | 734 (3.0) | 95.3% | 90.5 – 97.7 | 95.2% | 90.3 – 97.7 |
| Unvaccinated | 1818 (74.5) | 12863 (53.2) |  |  |  |  |
| **Outbreak in the facility / unit** | 628 (407) | 6280 |  |  |  |  |
| One-dose vaccinees (≥14 days) | 96 (28.7) | 3122 (59.3) | 76.6% | 70.0 – 81.8 | 73.0% | 65 – 79 |
| Two-doses vaccinees (≥7 days) | 4 (1.2) | 202 (3.8) | 85.8% | 57.2 – 95.3 | 85.0% | 55 – 95 |
| Unvaccinated | 235 (70.2) | 1940 (36.9) |  |  |  |  |
| **Systematic screening of HCWs** | 652 (373) | 6520 |  |  |  |  |
| One-dose vaccinees (≥14 days) | 91 (27.8) | 3251 (57.0) | 76.8% | 70.1 – 82.0 | 73.1% | 64.9 – 79.4 |
| Two-doses vaccinees (≥7 days) | 6 (1.8) | 362 (6.4) | 81.4% | 56.0 – 92.2 | 70.2% | 28.3 – 87.6 |
| Unvaccinated | 230 (70.3) | 2091 (36.7) |  |  |  |  |

^a^ Conditional logistic regression model adjusted for age, sex, job category, type of facility, health region and matching week

Abbreviations: CI=confidence interval; HCW=Healthcare worker; NE=not estimable; VE=vaccine effectiveness
